# A Novel Bleeding Risk Stratification Scheme in Japanese Patients with Non-valvular Atrial Fibrillation: The J-RISK AF study

**DOI:** 10.1101/2023.06.12.23291306

**Authors:** Masaharu Akao, Hirofumi Tomita, Michikazu Nakai, Eitaro Kodani, Shinya Suzuki, Kenshi Hayashi, Mitsuaki Sawano, Masahiko Goya, Takeshi Yamashita, Keiichi Fukuda, Toyonobu Tsuda, Mitsuaki Isobe, Kazunori Toyoda, Yoshihiro Miyamoto, Tomonori Okamura, Yusuke Sasahara, Ken Okumura, the J-RISK AF Research Group

## Abstract

**Background:** Oral anticoagulants (OAC) reduce the risk of ischemic stroke, but may increase the risk of major bleeding in non-valvular atrial fibrillation (NVAF) patients. Various risk scores, such as HAS-BLED, ATRIA, and ORBIT, have been proposed to assess the risk of major bleeding in NVAF patients receiving OAC. However, limited data are available on bleeding risk stratification in Japanese NVAF patients.

**Methods:** Of the 16,098 NVAF patients from the J-RISK AF study, the combined data of the five major AF registries in Japan (J-RHYTHM Registry, Fushimi AF Registry, Shinken Database, Keio interhospital Cardiovascular Studies, and Hokuriku-Plus AF Registry), we analyzed 11,539 patients receiving OAC (median age, 71 years; female, 39.6%; median CHADS_2_ score, 2). Multivariable Cox-hazard proportional analysis was performed to explore significant risk factors for major bleeding. Using those factors, we developed a novel bleeding risk stratification scheme and compared its predictive performance with previously reported risk scores.

**Results:** During the 2-year follow-up period, major bleeding occurred in 274 patients (1.3% per patient-year). On multivariable analysis, advanced age, uncontrolled hypertension, history of bleeding, anemia, thrombocytopenia, and concomitant antiplatelet agents were significantly associated with higher incidence of major bleeding. We developed a novel risk stratification system, J-RISK bleeding score, that had good predictive performance (C-statistics 0.67) for major bleeding. The predictive performance of our score was better than previous scores.

**Conclusion:** Our findings suggest that our novel risk stratification system, the J-RISK bleeding score, is more useful than previous score systems for Japanese NVAF patients receiving OAC.

## Clinical Perspective

### What is new?

- From the combined data of the five major registries of Japanese patients with non-valvular atrial fibrillation receiving oral anticoagulants, we identified significant risk factors for major bleeding.
- We developed a novel bleeding risk stratification scheme, the J-RISK bleeding score, and demonstrated that the predictive performance of this score was better than previously reported scores.

### What are the clinical implications?

- These risk factors and the novel score may be useful for identifying patients with non-valvular atrial fibrillation who are at risk for major bleeding.

## Introduction

Stroke prevention is an important issue for the management of patients with atrial fibrillation (AF) in clinical practice. Patients with AF are increasingly treated with oral anticoagulants (OAC), which have been reported to reduce the risk of thromboembolism and all-cause mortality. ^1, 2^ However, OAC may increase the risk of major bleeding in AF patients. The average annual incidences of fatal and major bleeding during warfarin therapy were reported to be 0.6% and 3.0%, respectively, approximately five times those expected without warfarin therapy. ^3^ Once major bleeding occurs, the case-fatality rate is high. A sub-analysis of the ARISTOTLE trial reported that mortality within 30 days occurred in 11.0% and 15.4% of patients receiving apixaban and warfarin, respectively. ^4^ Major bleeding is associated with increased risk of subsequent all-cause mortality and stroke/systemic embolism (SE) in the long-term among Japanese AF patients. ^5^

Various bleeding risk scores have been proposed in AF patients taking warfarin as an OAC. Most bleeding scores, such as HAS-BLED ^6^ and ATRIA ^7^, were developed and derived from studies conducted in the warfarin era. However, direct oral anticoagulants (DOACs) have been increasingly used for stroke prevention. Therefore, ORBIT ^8^ score is derived from data in AF patients taking either warfarin or DOAC, and has been proposed as alternative scores in the DOAC era. However, limited data are available on the clinical value of various bleeding risk scores among Japanese AF patients in contemporary clinical practice.

The aim of the present study was to determine risk factors for major bleeding, and to develop a novel scoring scheme for occurrence of major bleeding in a large-scale Japanese cohort of non-valvular atrial fibrillation (NVAF) patients who were administered OACs.

## Methods

### Study population

The detailed study design, patient enrollment, the definition of the measurements, and subjects’ baseline clinical characteristics of the J-RISK AF Research have been previously described. ^9^ We pooled the data from five major AF prospective registries in Japan: J-RHYTHM Registry (n=7,937), Fushimi AF Registry (n=3,749), Shinken Database (n=2,957), Keio Interhospital Cardiovascular Studies (n=783), and Hokuriku-Plus AF Registry (n=1,492). The data from each registry were collected and pooled in March 2016, and those from the Keio Interhospital Cardiovascular Studies were updated in April 2018. Of the 16,918 enrolled patients, 819 patients with valvular AF, 81 with lack of the event data of major bleeding, and 4,476 not receiving OAC at baseline were excluded. After excluding two patients with missing information about the type of OAC, analyses were finally performed on 11,539 patients. To balance the follow-up period among the registries, event data and follow-up period exceeding >730 days were excluded from the analysis. We collected demographics of the patients at baseline and follow-up data at 2 years.

This study was approved by the Ethics Committees of Hirosaki University Graduate School of Medicine (2015-117, 2017-1051), National Cerebral and Cardiovascular Center (M27-092-4), National Hospital Organization Kyoto Medical Center (15-101), Cardiovascular Institute (279), Kanazawa University Graduate School of Medical Science (2035-1, 2460-1), and Keio University School of Medicine (20120029), and was performed within Ethics Committee-approved research protocols at other institutes.

### Definitions

The primary endpoint in the analysis was the incidence of major bleeding. The definition of major bleeding varies among the registries but are in accordance with the criteria of the International Society on Thrombosis and Haemostasis (ISTH), which consist of the following: (i) reduction in hemoglobin level of at least 2 g/dl; (ii) transfusion of at least 2 units of blood; and (iii) symptomatic bleeding at a critical area or organ (intracranial, intraocular, intraspinal, intraarticular, intramuscular with compartment syndrome, pericardial, retroperitoneal). Major bleeding was defined as: (i) intracranial hemorrhage, gastrointestinal hemorrhage, and other hemorrhages requiring hospitalization in the J-RHYTHM Registry; (ii) according to the ISTH criteria in the Fushimi AF Registry and the Keio Interhospital Cardiovascular Studies; (iii) bleeding that required emergency hospitalization which includes intracranial and extracranial hemorrhage in the Shinken Database; and (iv) intracranial hemorrhage, including hemorrhagic stroke, hemorrhagic events requiring transfusion, and hemorrhagic events with reduction of hemoglobin by >2 g/dl, in the Hokuriku-Plus AF Registry.

Valvular AF was defined as AF with a rheumatic mitral stenosis or a prosthetic heart valve. OAC included warfarin and DOAC (dabigatran, rivaroxaban, apixaban, and edoxaban). Japanese treatment guidelines during this study period recommended different target prothrombin time international normalized ratios (PT-INRs) for patients taking warfarin: 1.6-2.6 for elderly patients (≥70 years old) and 2.0-3.0 for younger patients (<70 years old). ^10^ Antiplatelet agents included aspirin and thienopyridines.

We categorized the patients in relation to the risk of major bleeding, using the HAS-BLED score ^6^ (uncontrolled Hypertension, Abnormal renal/liver function, Stroke, Bleeding history or predisposition, Labile PT-INR, Elderly, and Drug/alcohol concomitantly), ATRIA score ^7^ (Anemia, Renal disease, Age, Prior bleeding, and Hypertension), and ORBIT score ^8^ (Older age, Reduced hemoglobin or hematocrit, Bleeding history, Insufficient kidney function, and Treatment with antiplatelets), as originally described, except for the factor ‘L’ (labile PT-INR) of the HAS-BLED score. For the labile PT-INR criterion, we defined the value of PT-INR as over 2.6 for elderly patients (≥70 years old) or 3.0 for younger patients (<70 years old)) at baseline according to the Japanese AF guidelines, ^10^ because the values of PT-INR were collected at the time of enrollment. Furthermore, for warfarin control, we defined appropriate as PT-INR within target range, overdose as that over target range, and underdose as that under target range at baseline.

### Statistical analysis

Categorical variables are presented as numbers and percentages. Continuous variables are presented as mean and standard deviation for normally distributed data, or median and interquartile range for non-normal distribution. We compared categorical variables using chi-square test and continuous variables using independent samples t-test for normally distributed data or Mann-Whitney U-test for non-normal distribution. The cut-off of continuous variables was based on previous studies. Crude event rates were presented by % per patient-year. We carried out univariable Cox proportional hazards models after confirming proportional hazards, and constructed multivariable Cox proportional hazards models to determine the variables associated with the incidence of major bleeding. All variables were evaluated for collinearity. The candidate variables for Model 1 were chosen from the significant variables on the univariable Cox proportional hazards models. Those for the Model 2 were the significant variables included in Model 1. We used multiple imputation method for missing data (6.3% of total data). Multiple imputation method using chained equation algorithm was conducted based on 10 replications, assuming missing at random mechanism. Imputed variables are shown in Supplemental Table 1. A score was assigned to each significant risk factor based on the coefficient calculated by logarithmically transformed hazard ratios (HRs). We assessed model performance by examining calibration and discrimination at 2 years of follow-up in the J-RISK AF cohort. Calibration was evaluated by plotting event rates of major bleeding according to J-RISK bleeding score and calibration plot in the J-RISK AF cohort. Confidence intervals of event rates were calculated by bootstrap resampling of 1,000 replications. Modified Hosmer-Lemeshow test was performed to determine the goodness-of-fit of the J-RISK bleeding score. Discrimination was evaluated using C-Statistics, for comparison with previous risk stratification schemes such as HAS-BLED, ATRIA, and ORBIT scores. In addition, a sensitivity analysis was performed in patients divided into subgroups consisting of those taking warfarin and those taking DOAC. Significance was set at a two-sided p-value of <0.05. Analyses were performed using JMP version 13.2.0 (SAS Institute, Cary, NC) and R version 4.1.0 for Windows.

## Results

Of 11,539 NVAF patients, major bleeding occurred in 274 patients during the 2-year follow-up period (1.3% per patient-year); 238 patients were taking warfarin (1.4% per patient-year); and 36 patients were taking DOAC (1.3% per patient-year) (p=0.697). Intracranial hemorrhage occurred in 93 of 274 patients with major bleeding (33.9%) (0.4% per patient-year).

Demographics of the patients at baseline, categorized by occurrence of major bleeding during follow-up period, are shown in Table 1. NVAF patients with major bleeding were significantly older and had lower body weight, higher systolic blood pressure (SBP), lower hemoglobin level, and lower platelet count. They were significantly more likely to have history of stroke/transient ischemic attack (TIA), heart failure, hypertension, coronary artery disease, chronic kidney disease, and history of bleeding, than those without major bleeding. Baseline stroke risk scores (CHADS_2_ and CHA_2_DS_2_-VASc) and bleeding risk scores (HAS-BLED, ATRIA, and ORBIT) were significantly higher in patients with major bleeding.

**Table 1.**
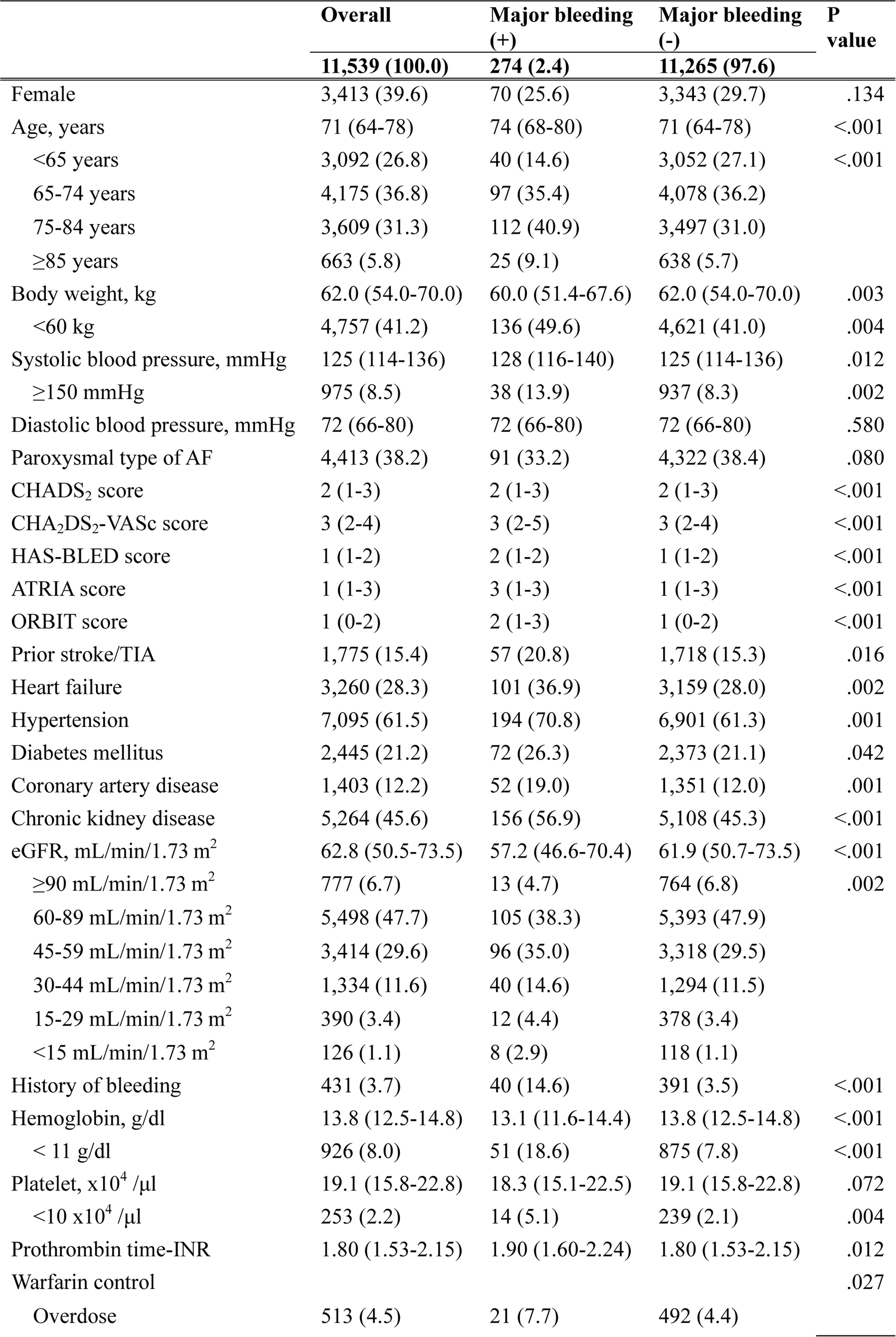
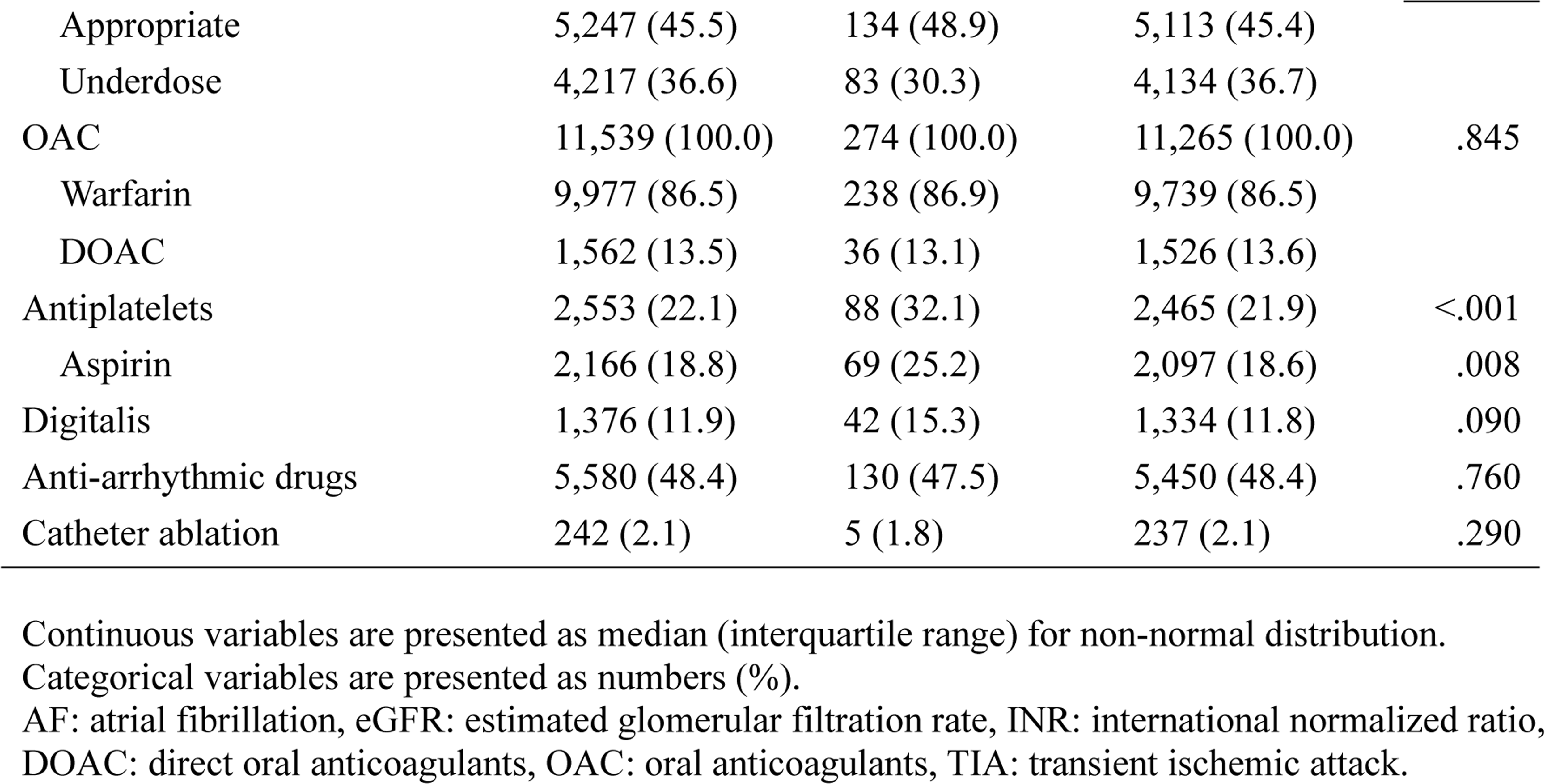
Patient characteristics, medication and previous therapy at baseline

Type of administrated OAC was comparable between NVAF patients with and without major bleeding at baseline. Concomitant use of antiplatelets, predominantly aspirin, was found more often in major bleeding patients. PT-INR at baseline was significantly higher in patients with major bleeding. Warfarin was appropriately controlled in 45.5% of patients, but NVAF patients with major bleeding were more often controlled at overdoses and less often at underdoses of warfarin.

### Factors associated with incidence of major bleeding and score assignment

We performed Cox proportional hazards analysis to identify variables associated with incidence of major bleeding (Table 2). On an univariable analysis, age 65-74 and ≥75 years, body weight <60 kg, SBP ≥150 mmHg, history of stroke/TIA, heart failure, hypertension, diabetes mellitus, coronary artery disease, history of bleeding, hemoglobin <11 g/dl, platelet <10×10^4^ /μl, eGFR <15 mL/min/1.73 m^2^, and concomitant use of antiplatelets were significant. On a multivariable model, which included factors that were significantly associated in the univariable model (Model 1), we indicated that age 65-74 and ≥75 years, SBP ≥150 mmHg, history of bleeding, hemoglobin <11 g/dl, platelet <10×10^4^/μl, and concomitant use of antiplatelets were significantly associated with higher incidence of major bleeding. Multivariable Model 2 was constructed by excluding non-significant variables on Model 1. No collinearity was found for any of the variables used in the multivariable models (variance inflation factor <10).

**Table 2.** Cox proportional hazards analysis for variables associated with incidence of major bleeding

|  | Univariable |  | Multivariable: Model 1 |  | Multivariable: Model 2 |  | Coefficient | Points |
| --- | --- | --- | --- | --- | --- | --- | --- | --- |
|  | HR | 95% CI | HR | 95% CI | HR | 95% CI |  |  |
| Age |  |  |  |  |  |  |  |  |
| <65 years | Reference |  | Reference |  | Reference |  |  |  |
| 65-74 years | 1.79 | 1.24, 2.61 | 1.66 | 1.06, 2.59 | 1.87 | 1.21, 2.86 | 0.62 | 1 |
| ≥75 years | 2.52 | 1.79, 3.63 | 1.81 | 1.16, 2.83 | 2.00 | 1.31, 3.06 | 0.69 | 2 |
| Body weight <60 kg | 1.42 | 1.10, 1.82 | 1.07 | 0.80, 1.43 |  |  |  |  |
| Systolic blood pressure ≥150 mmHg | 1.70 | 1.15, 2.59 | 1.70 | 1.14, 2.54 | 1.74 | 1.19, 2.55 | 0.55 | 1 |
| History of stroke/TIA | 1.48 | 1.10, 1.97 | 1.18 | 0.84, 1.65 |  |  |  |  |
| Heart failure | 1.49 | 1.16, 1.90 | 1.16 | 0.87, 1.55 |  |  |  |  |
| Hypertension | 1.52 | 1.18, 1.99 | 1.17 | 0.86, 1.58 |  |  |  |  |
| Diabetes mellitus | 1.34 | 1.02, 1.74 | 1.27 | 0.93, 1.73 |  |  |  |  |
| Coronary artery disease | 1.73 | 1.27, 2.32 | 1.12 | 0.76, 1.66 |  |  |  |  |
| History of bleeding | 4.72 | 3.31, 6.55 | 3.95 | 2.70, 5.78 | 4.21 | 2.93, 6.05 | 1.43 | 3 |
| Hemoglobin <11 g/dl | 3.06 | 2.21, 4.14 | 1.98 | 1.35, 2.89 | 2.15 | 1.52, 3.04 | 0.76 | 2 |
| Platelet <10 x10<sup>4</sup> /μl | 2.95 | 1.64, 4.87 | 2.58 | 1.49, 4.49 | 2.51 | 1.45, 4.34 | 0.92 | 2 |
| eGFR <15 mL/min/1.73 m<sup>2</sup> | 3.09 | 1.40, 5.83 | 1.60 | 0.75, 3.39 |  |  |  |  |
| Warfarin (vs. DOAC) | 1.06 | 0.74, 1.49 |  |  |  |  |  |  |
| Warfarin control (vs. DOAC) |  |  |  |  |  |  |  |  |
| Overdose | 1.56 | 0.99, 2.67 |  |  |  |  |  |  |
| Appropriate | 0.96 | 0.66, 1.40 |  |  |  |  |  |  |
| Underdose | 0.75 | 0.58, 1.06 |  |  |  |  |  |  |
| Antiplatelets | 1.67 | 1.29, 2.15 | 1.36 | 1.00, 1.89 | 1.56 | 1.18, 2.06 | 0.44 | 1 |

CI: confidence interval, eGFR: estimated glomerular filtration rate, HR: hazard ratio, DOAC: direct oral anticoagulants, TIA: transient ischemic attack.

Based on this multivariable model’s regression coefficients, we propose a new score (“J-RISK bleeding score”): history of bleeding is assigned 3 points; age ≥75 years, hemoglobin <11 g/dl, and platelet <10×10^4^ /μl are 2 points; and age 65-74 years, SBP ≥150 mmHg, and concomitant use of antiplatelets are 1 point each, resulting in a risk scheme with a possible range of 0 to 11 points.

### Model performance

Figure 1 shows the crude incidences of major bleeding stratified by J-RISK bleeding score. Observed major bleeding rates increased with increasing risk score in the entire cohort (Figure 1A) and patients were divided into subgroups consisting of those receiving warfarin or DOAC (Figure 1B). Event rates calculated by the bootstrap method were similar to the observed crude incidence rates (Supplemental Table 2). Calibration plots comparing the predicted major bleeding rates by our model and those actually observed at 2 years of follow-up are displayed in Figure 2. In the overall patients (Figure 2A) the predicted event rates by the J-RISK bleeding score were a good fit for actual event rates (modified Hosmer-Lemeshow test; p=0.965). Also, in the patients divided into the subgroups of those receiving warfarin or DOAC (Figure 2B), the J-RISK bleeding score showed good calibration for major bleeding rates (modified Hosmer-Lemeshow test; p=0.876 and p=0.998, respectively).

**Figure 1.**
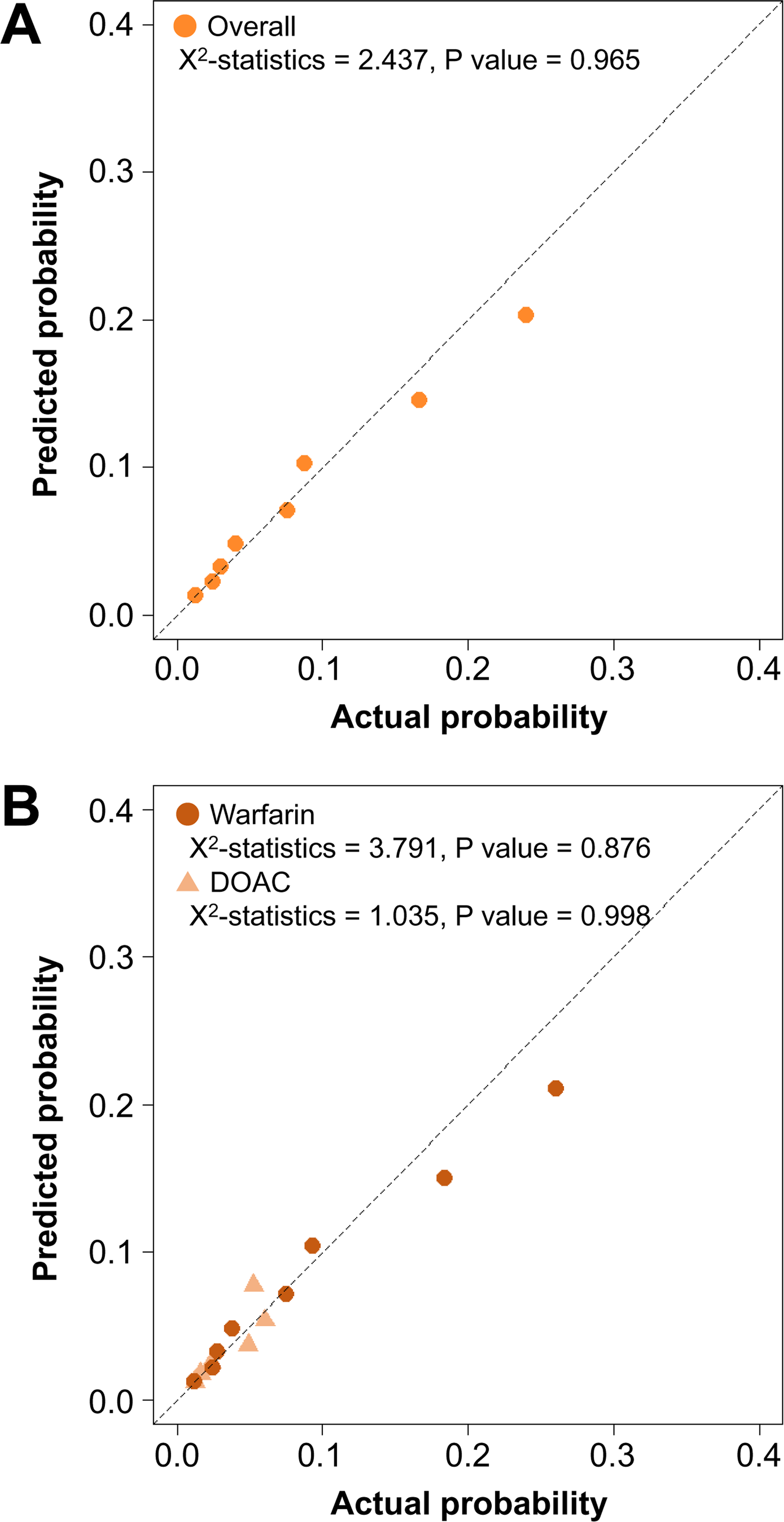
Crude incidence rate of major bleeding, stratified by the J-RISK bleeding score in the overall patients (A) and the patients with warfarin or DOAC (B) during follow-up period. The number over the bar presents the crude incidence rate (% per patient-year) of the J-RISK bleeding score. DOAC: direct oral anticoagulants. SBP: systolic blood pressure.

**Figure 2.**
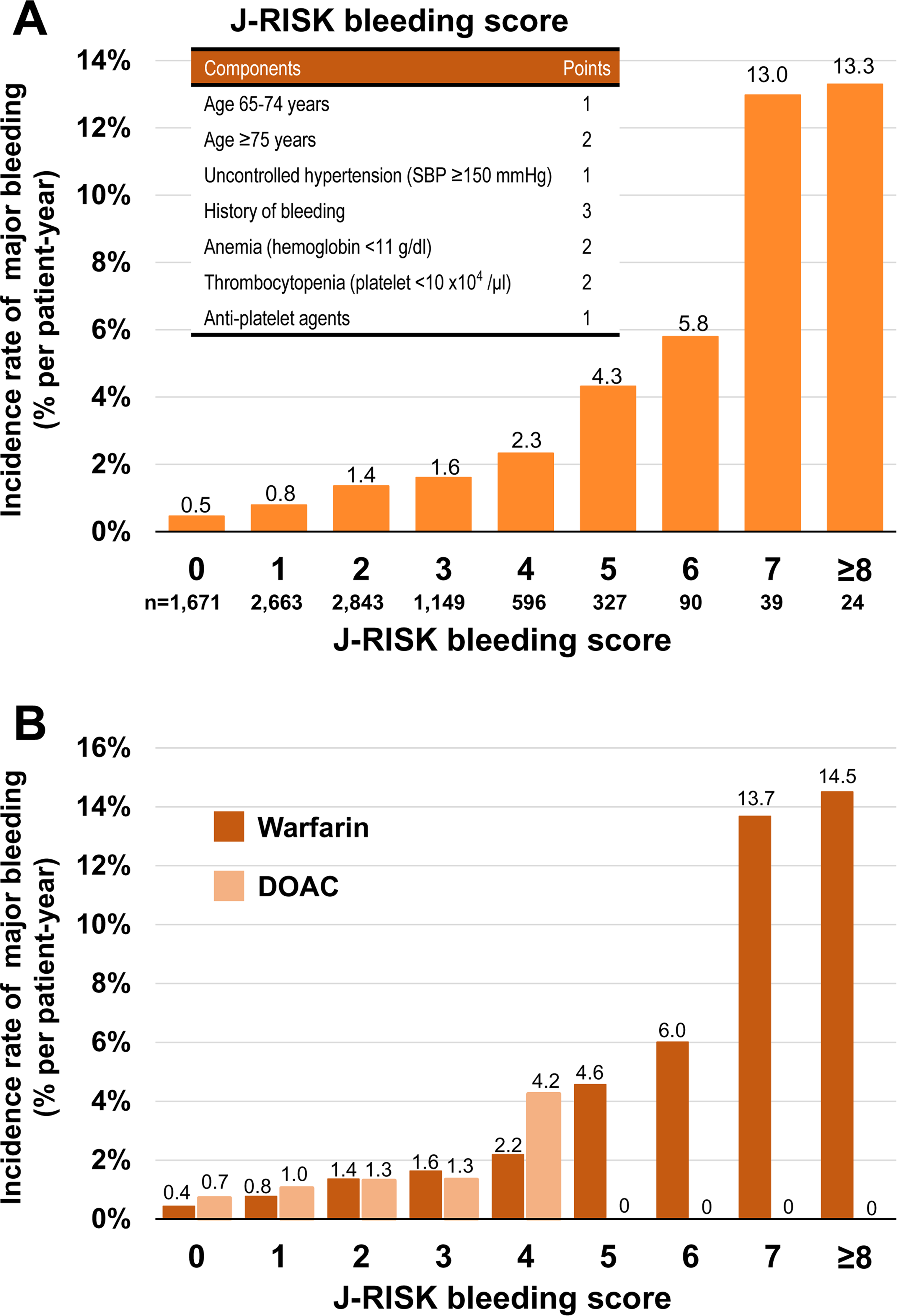
Calibration plots of actual outcome versus predictions for the J-RISK bleeding score in the overall patients (A) and the patients with warfarin or DOAC (B) at 2 years of follow-up. The modified Hosmer-Lemeshow test was performed to determine the goodness-of-fit of the J-RISK bleeding score. Chi-squared statistics and p-value are presented. DOAC: direct oral anticoagulants.

C-statistics for major bleeding are shown in Table 3. The J-RISK bleeding score had significantly higher C-statistics than HAS-BLED and ATRIA scores; furthermore, it was comparable to the ORBIT score. In a sensitivity analysis, J-RISK bleeding score also showed the best discrimination in patients taking warfarin on C-statistics. On the other hand, in patients taking DOAC, C-statistics of the J-RISK bleeding score were similar to those of other schemes (Table 3).

**Table 3.** Comparison of bleeding risk schemes

| **Risk scheme** | **C-Statistics** | **95% CI** | **P value** |
| --- | --- | --- | --- |
| **Overall** |  |  |  |
| J-RISK bleeding score | 0.67 | 0.63, 0.70 |  |
| HAS-BLED score | 0.60 | 0.57, 0.64 | <.01 |
| ATRIA score | 0.63 | 0.60, 0.66 | .01 |
| ORBIT score | 0.65 | 0.62, 0.68 | .22 |
| **Warfarin** |  |  |  |
| J-RISK bleeding score | 0.67 | 0.64, 0.70 |  |
| HAS-BLED score | 0.61 | 0.57, 0.65 | <.01 |
| ATRIA score | 0.64 | 0.60, 0.67 | .01 |
| ORBIT score | 0.66 | 0.62, 0.69 | .23 |
| **DOAC** |  |  |  |
| J-RISK bleeding score | 0.64 | 0.55, 0.74 |  |
| HAS-BLED score | 0.62 | 0.55, 0.71 | .70 |
| ATRIA score | 0.61 | 0.51, 0.70 | .30 |
| ORBIT score | 0.64 | 0.55, 0.72 | .71 |

CI: confidence interval, DOAC: direct oral anticoagulants

## Discussion

The principal findings of this study are as follows: (i) the incidence rate of major bleeding was 1.3% per patient-year in AF patients receiving OAC; (ii) on a multivariable model, age 65-74 and ≥ 75 years, SBP ≥ 150 mmHg, history of bleeding, hemoglobin <11 g/dl, platelet <10×10^4^ /μl, and concomitant use of antiplatelets were significantly associated with higher incidence of major bleeding; and (iii) the novel J-RISK bleeding score had better predictive performance than previous bleeding risk scores such as HAS-BLED, ATRIA, and ORBIT.

In the global phase III randomized clinical trials of DOAC, the annualized rate of major bleeding was approximately 3% in AF patients taking warfarin, and ranged from 2.1% to 3.6% in those taking DOAC. ^2^ In a sub-analysis of these trials of DOAC in Japanese AF patients, the incidence rate of major bleeding ranged from 3.3%/year to 4.0%/year in those with taking warfarin, and from 1.2%/year to 5.5%/year in those taking DOAC. ^11–14^ In previous observational studies, incidence rates of major bleeding were 1.56% per patient-year in the Euro Heart Survey on Atrial Fibrillation, ^6^ and 1.9% per patient-year in the Swedish Atrial Fibrillation cohort study, ^15^ which were comparable to the present study.

### Clinical factors associated with major bleeding

Advanced age is one of the most consistent factors associated with major bleeding in the previous studies, and is included in all bleeding scores, such as HAS-BLED, ATRIA, and ORBIT, with an age cut-off of 65 years or 75 years. The ATRIA, and ORBIT scores included an age cut-off of 75 years, whereas the HAS-BLED score and our score included an age cut-off of 65 years. In the present analysis, adjusted HR in patients of age ≥75 years was modestly higher than those of age 65-74 years, but scores of these age ≥75 years were assigned 2 points, based on their coefficients rounded to the nearest integer.

Concerning hypertension, uncontrolled hypertension but not history of hypertension, is included in the HAS-BLED, and J-RISK bleeding scores, whereas the ATRIA score includes history of hypertension. ‘Uncontrolled hypertension’ is defined as SBP >160 mmHg in the HAS-BLED score, and ≥150 mmHg in our score. The cut-off of SBP value of 150 mmHg was adopted based on our previous studies in Japan. ^16 17^

Anemia, defined as hemoglobin level <11 g/dl, was significantly associated with major bleeding in this study, which is consistent with previous scores. The ATRIA, and ORBIT scores include anemia; however, the definition of anemia differs for each score. In the present score, we determined the cut-off of hemoglobin level <11 g/dl, based on moderate or severe anemia, using the World Health Organization classification of anemia.

Concomitant antiplatelets were significantly associated with a higher incidence of major bleeding in the present study, and are included in the HAS-BLED, ORBIT, and J-RISK bleeding scores. The AFIRE trial reported that rivaroxaban monotherapy was non-inferior to combination therapy of rivaroxaban and antiplatelets for thromboembolism and was superior for major bleeding in AF patients with stable coronary artery disease. ^18^ Physicians are encouraged to withhold concomitant antiplatelet therapy, taking thromboembolic and bleeding risk factors for each patient into consideration.

In the present study, thrombocytopenia, defined as platelet count (<10×10^4^/μl) was significantly associated with a higher incidence of major bleeding. This result is consistent with the previous study from Korea, which showed that lower platelet counts were associated with a higher risk of bleeding events. ^19^ Although a sub-analysis of the J-RHYTHM Registry reported that the association between low platelet count (<10×10^4^/μl) and major bleeding was not significant (p=0.073) in a multivariable model ^20^, the present pooled analysis, which included the J-RHYTHM Registry, found a significant association.

### Comparison with previous scores

The components of the J-RISK bleeding score overlap with those of other scores such as HAS-BLED, ATRIA, and ORBIT. Our findings suggest that the J-RISK bleeding score outperformed the HAS-BLED, ATRIA, and ORBIT scores; the C-statistics of the J-RISK bleeding score (0.67, 0.67, and 0.64 in the total cohort, the warfarin subgroup, and the DOAC subgroup, respectively) indicated relatively good performance. O’Brien et al. demonstrated that the C-statistics of the ORBIT score were 0.67 in the ORBIT-AF cohort and 0.62 in the ROCKET-AF cohort. ^8^ In addition, they reported that C-statistics of the HAS-BLED and ATRIA scores were approximately 0.6 in both the ORBIT-AF and ROCKET-AF cohorts. Furthermore, Lip et al. reported that C-statistics for ATRIA, HAS-BLED, and ORBIT scores were 0.59, 0.58, and 0.61, respectively, using data from a Danish nationwide database. ^21^ Most of the previous scores included reduced renal function as an independent risk factor, whereas our score did not. Kidney function may dynamically change and worsen with age and comorbidities. ^22^ The J-RISK bleeding score may be a useful predictive score for major bleeding, which includes well-known and easily available risk factors for major bleeding and elements that are relevant to all patients taking OAC in the contemporary population of AF. Physicians are encouraged to modify the reversible factors, such as anemia, blood pressure, and unnecessary concomitant antiplatelets, in daily practice.

### Limitations

There are several limitations to this study. The present results were not prospectively validated by external data. Medications and indications for OAC were selected at the discretion of the attending physician. We also have no data on time in therapeutic range for individual patients taking warfarin. We collected prescription data at baseline; however, we do not know the date of interruption or resumption of drugs including OAC. In addition, we do not know exact type of OAC at the time of major bleeding. Bleeding risk is dynamic and may change with age and incident risk factors or by mitigation of modifiable risk factors. ^23, 24^

## Conclusion

We demonstrated that advanced age, uncontrolled hypertension, history of bleeding, anemia, thrombocytopenia, and concomitant use of antiplatelets were significantly associated with higher incidence of major bleeding. We developed a novel risk stratification system, J-RISK bleeding score, for the incidence of major bleeding in Japanese NVAF patients taking OAC. This score may be more useful than previous score systems in Japanese NVAF patients.

## Acknowledgements

J-RISK AF Research Group Investigators/Collaborators Pooled analysis for identifying RISK factors for Atrial Fibrillation and cardioembolic stroke in the Japanese population (J-RISK AF Research group) is composed of the following investigators: Chief Investigator: Ken Okumura (Hirosaki University, Saiseikai Kumamoto Hospital); Assistant Chief Investigator: Hirofumi Tomita (Hirosaki University); Participating Investigators: Toshiharu Ninomiya, Jun Hata (Kyushu University); Hiroyasu Iso, Hironori Imano, Isao Muraki, Cui Renzhe (Osaka University); Yoshihiro Miyamoto, Yoshihiro Kokubo, Michikazu Nakai, Yusuke Sasahara, Kazunori Toyoda, Manabu Inoue, Masahito Takagi (National Cerebral and Cardiovascular Center); Kazutaka Aonuma, Nobuyuki Murakoshi (University of Tsukuba); Gen Kobashi, Toshimi Sairenchi (Dokkyo Medical University); Takeshi Yamashita, Shinya Suzuki (The Cardiovascular Institute); Masaharu Akao (National Hospital Organization Kyoto Medical Center); Keiichi Fukuda, Tomonori Okamura, Hiroaki Miyata, Mitsuaki Sawano, Norimichi Hirahara, Daisuke Sugiyama (Keio University); Kenshi Hayashi, Hiroshi Furusho, Toyonobu Tsuda (Kanazawa University); Mitsuaki Isobe (Tokyo Medical and Dental University, Sakakibara Heart Institute); Tetsushi Furukawa, Masahiko Goya (Tokyo Medical and Dental University); Eitaro Kodani (Nippon Medical School Tama-Nagayama Hospital); Yoshifusa Aizawa (Tachikawa Medical Center); Fujiko Irie (Ibaraki Prefectural Government); Kohei Tanizaki (Sakakibara Heart Institute).

## Data Availability Statements

The data underlying this article will be shared on reasonable request to the corresponding author.

